# Sputnik Light booster after Sputnik V vaccination induces robust neutralizing antibody response to B.1.1.529 (Omicron) SARS-CoV-2 variant

**DOI:** 10.1101/2021.12.17.21267976

**Authors:** IV Dolzhikova, AA Iliukhina, AV Kovyrshina, AV Kuzina, VA Gushchin, AE Siniavin, AA Pochtovyi, EV Shidlovskaya, NA Kuznetsova, MM Megeryan, AS Dzharullaeva, AS Erokhova, FM Izhaeva, DM Grousova, AG Botikov, DV Shcheblyakov, AI Tukhvatulin, OV Zubkova, DY Logunov, AL Gintsburg

## Abstract

COVID-19 vaccination campaign has been launched around the world. More than 8 billion vaccines doses have been administered, according to the WHO. Published studies shows that vaccination reduces the number of COVID-19 cases and dramatically reduces COVID-19-associated hospitalizations and deaths worldwide. In turn, the emergence of SARS-CoV-2 variants of concern (VOC) with mutations in the receptor-binding domain (RBD) of S glycoprotein poses risks of diminishing the effectiveness of the vaccination campaign. In November 2021, the first information appeared about a new variant of the SARS-CoV-2 virus, which was named Omicron. The Omicron variant is of concern because it contains a large number of mutations, especially in the S glycoprotein (16 mutation in RBD), which could be associated with resistance to neutralizing antibodies (NtAB) and significantly reduce the effectiveness of COVID-19 vaccines. Neutralizing antibodies are one of the important parameters characterizing the protective properties of a vaccine. We conducted a study of neutralizing antibodies in the blood serum of people vaccinated with Sputnik V, as well as those revaccinated with Sputnik Light after Sputnik V. Results showed that a decrease in the level of neutralizing antibodies was observed against SARS-CoV-2 Omicron (B.1.1.529) variant in comparison to B.1.1.1 variant. Analysis of the sera of individuals vaccinated with Sputnik V 6-12 months ago showed that there was a decrease in the level of neutralizing antibodies by 11.76 folds. While no direct comparison with other vaccines declines has been done in this study, we note their reported decline in antibody neutralization at a much more significant level of 40-84 times. At the same time, the analysis of sera of individuals who were vaccinated with Sputnik V, and then revaccinated Sputnik Light, showed that 2-3 months after revaccination the decrease in the level of neutralizing antibodies against the Omicron variant was 7.13 folds. Despite the decrease in NtAb, we showed that all revaccinated individuals had NtAb to Omicron variant. Moreover, the NtAb level to Omicron variant in revaccinated sera are slightly higher than NtAb to B.1.1.1 in vaccinated sera.

## Introduction

A new variant of SARS-CoV-2 B.1.1.529 (Omicron) presumably appeared in Botswana. It spreads faster than delta (B.1.617.2) and beta (B.1.351) variants. As of December 15, 2021, according to the GISAID database, the largest number of people infected with this variant of the coronavirus are in the UK and South Africa [1].

It was found that Omicron variant contains 60 mutations relative to variant B.1, of which more than 30 were detected in the S glycoprotein. Multiple mutations in receptor-binding domain (RBD) and the N-terminal domain of S glycoprotein are associated with resistance to neutralizing antibodies (NtAb). Omicron variant has a large number of rare and unique mutations. Such rapid evolution could have occurred in a population of immunocompromised individuals who have low numbers of CD4+ T cells and the virus could establish a persistent infection because of a lack of killer-T-cell responses [2].

Due to the huge number of mutations in the S glycoprotein of the SARS-CoV-2 B.1.1.529 (Omicron) variant, there are concerns that this variant will successfully evade neutralizing antibodies in convalescent and vaccinated individuals, which could significantly reduce the effectiveness of vaccines and monoclonal antibodies used for prevention and treatment of COVID-19.

It was shown that sera from vaccinated individuals neutralized the B.1.1.529 variant to a much lesser extent than any other analyzed SARS-CoV-2 variant, individuals were vaccinated with Moderna, ChAdOx1 (Vaxzevria, AstraZeneca), BNT162b2 (Comirnaty, Pfizer/BioNTech) and ChAdOx1 prime/BNT162b2 boost [3]. The neutralization activity against B.1.1.529 was higher in sera from superimmune individuals with hybrid immunity (infected and vaccinated or vaccinated and infected) [3-5]. In conditions of decreasing virus neutralizing activity of antibodies, T-cell mediated immunity becomes an essential barrier to prevent severe COVID-19 [5]. COVID-19 vaccines that can elicit post-vaccinal T-cell immunity should be studied.

## Results

We performed analysis of neutralization antibody in sera samples of individuals vaccinated with recombinant adenoviral-based vaccine Sputnik V (rAd26-S + rAd5-S) and revaccinated with Sputnik Light vaccine (rAd26-S). We isolated the viable SARS-CoV-2 virus Omicron variant (B.1.1.529) from a patient who arrived from South Africa (Figure 1). The virus was genetically characterized (hCoV-19/Russia/MOW-Moscow_PMVL-O16/2021) and used to analyze the virus-neutralizing activity of the sera of vaccinated and revaccinated individuals. The study was conducted with sera from people who were vaccinated with Sputnik V 6-12 months ago, as well as sera from people who were revaccinated with Sputnik Light 2-3 months ago (Table S1). RBD-specific IgG was detected in all analyzed sera samples (Figure 2).

**Figure 1.**
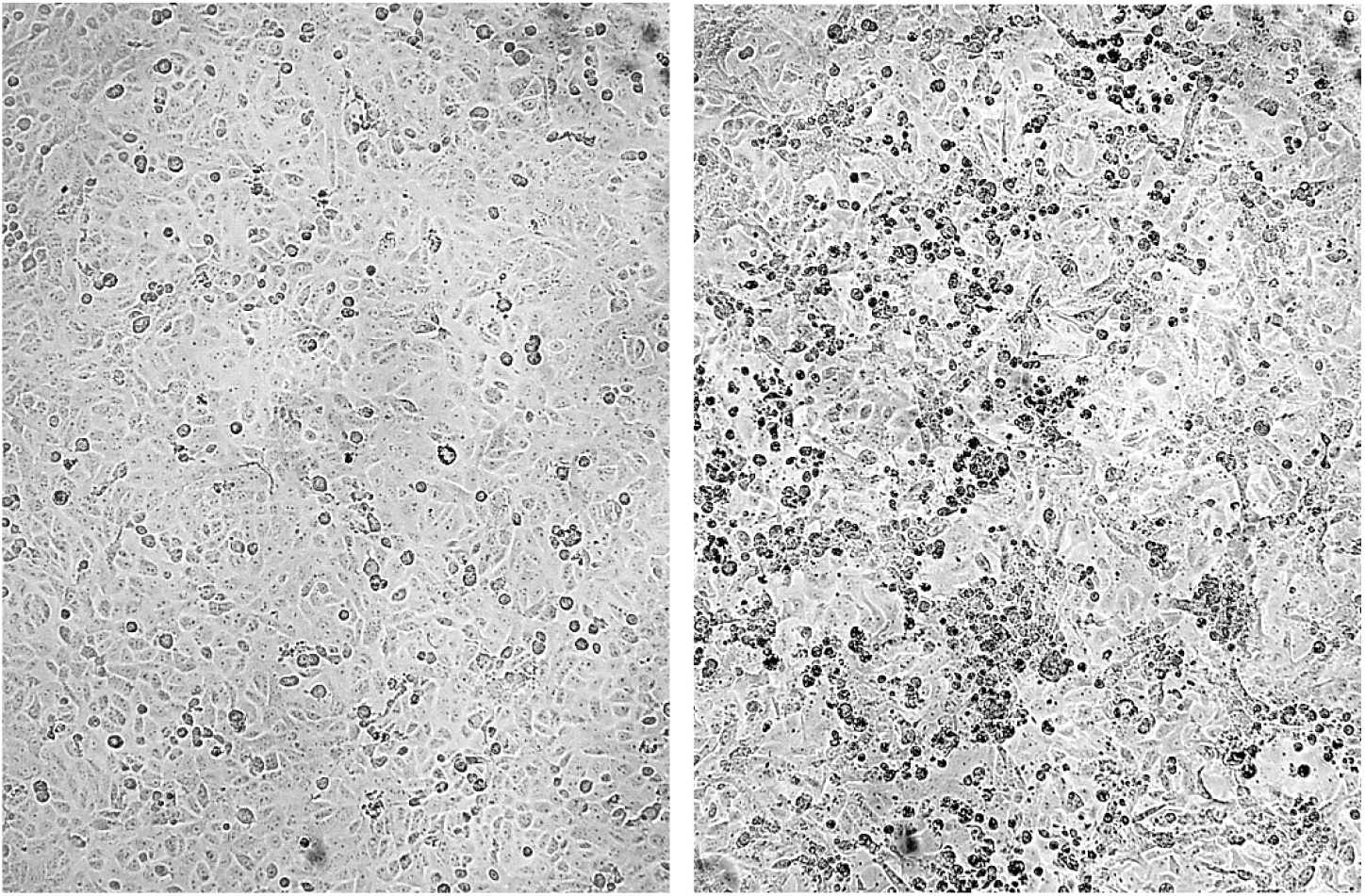
Photographs of Vero E6 cells: left - control cells, right - cells infected with B.1.1.529.

**Figure 2.**
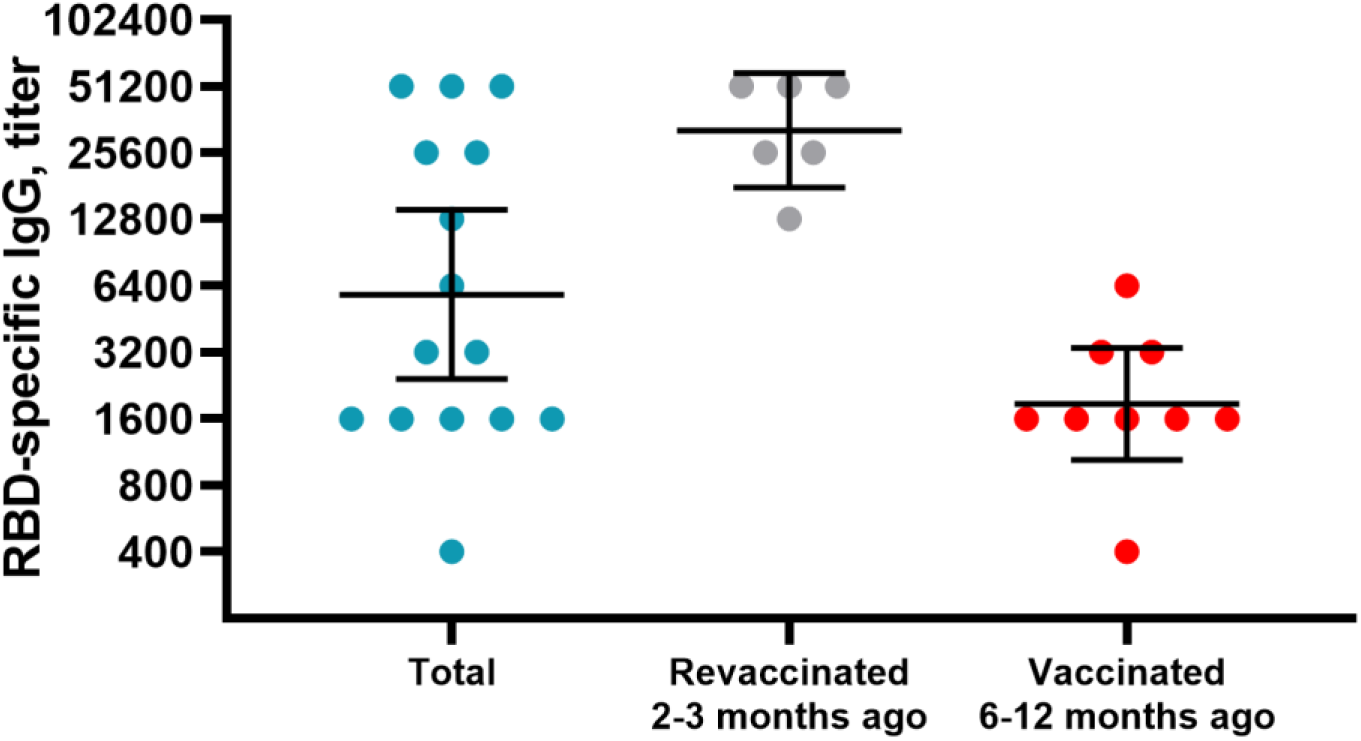
Titers of RBD-specific IgG in blood sera samples of individuals vaccinated with Sputnik V and revaccinated with Sputnik Light. The figure shows individual data for the studied sera, geometric mean and 95% confidence interval.

Analyzing NtAb in the total pool of sera, we detected a decrease to the Omicron variant by 9.62 folds (Figure 3) in comparison with the variant B.1.1.1. The NtAb in sera of people vaccinated with Sputnik V 6-12 months ago decreased in relation to the Omicron variant by 11.76 folds. Notably, when analyzing the sera of people revaccinated with Sputnik Light after Sputnik V, we showed a smaller decrease in NtAb to the Omicron variant - 7.13 folds. It is important to note that after revaccination with Sputnik Light, despite the NtAb decrease, NtAb were detected in the serum of all revaccinated individuals. Moreover, the level of NtAb to Omicron variant in sera of revaccinated with Sputnik Light were statistically same (p=0.2517, Mann-Whitney test) with NtAb to B.1.1.1 in sera of vaccinated with Sputnik V.

**Figure 3.**
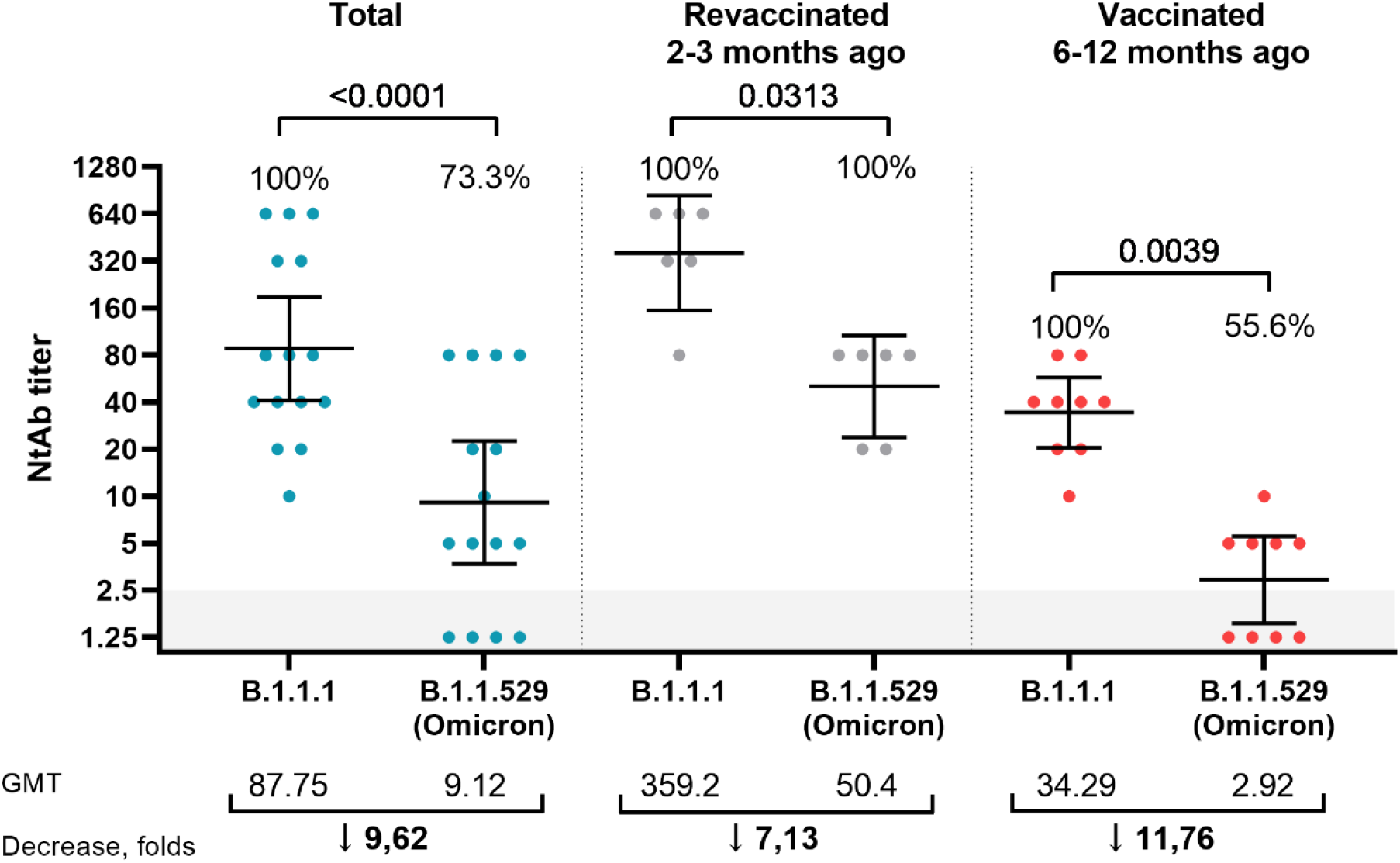
Titers of neutralizing antibodies in blood sera samples of individuals vaccinated with Sputnik V and revaccinated with Sputnik Light. The figure shows individual data for the studied sera, geometric mean and 95% confidence interval. The figure shows the p-value (Wilcoxon test), % of individuals with detectable NtAb to the SARS-CoV-2, geometric mean and the level of NtAb reduction. Grey box – limit of detection. Values below the limit of detection were assigned a value of NtAb 1.25.

## Discussion

Our data show that vaccination with Sputnik V and revaccination with Sputnik Light allows to induce robust neutralizing antibody response. We see that the NtAb decrease against the Omicron variant is the most significant than against the other SARS-CoV-2 variants of concern (VOCs). In our previous studies, we showed the change in the serum NtAb level of Sputnik V vaccinated against SARS-CoV-2 VOCs (Alfa B.1.1.7, Beta B.1.351, Gamma B.1.1.28/P.1, Delta B.1.617.2), which was most significant for the Beta variant [6]. The Omicron variant carries a huge number of mutations, which led to NtAb decrease. Remarkably, despite the decrease in NtAb, we showed that among individuals who were revaccinated with Sputnik Light after vaccination with Sputnik V, NtAb were detected in all blood serum samples in levels statistically same with those detected in sera of Sputnik V vaccinated to B.1.1.1 variant. So, the obtained data indicate the need for revaccination to maintain a high level of antibodies neutralizing SARS-CoV-2 virus.

This study shows a preliminary data on neutralizing activity of Sputnik V-vaccinated and Sputnik Light revaccinated sera to B.1.1.529 (Omicron) SARS-CoV-2 variant. This study has several limitations: 1) the relatively small number of tested samples, which may not reflect the range of neutralizing antibodies in different groups of vaccinated, 2) limited data on the dynamics over time of neutralizing antibodies to the Omicron variant after vaccination and revaccination. Additional work is currently underway to study the level of neutralizing antibodies in expanded sample sizes, as well as to study the effectiveness of the Sputnik V vaccine against Omicron variant in a lethal model in hACE2-transgenic mice.

Further studies are important to understand the effectiveness of COVID-19 vaccines against the new SARS-CoV-2 Omicron variant. If there will be a critical decrease in effectiveness (lower than recommended by WHO), it is necessary to consider change the antigenic composition of the vaccines. In this case, the efficacy of the new vaccine candidate should be studied against Omicron, as well as Delta, the proportion of which remains prevalent.

## Materials and methods

### Ethics statement

The study was conducted according to the guidelines of the Declaration of Helsinki, and approved by the Local Ethics Committee of Gamaleya NRCEM (Protocol No. 17 of December 03, 2021).

### Sera samples

Peripheral blood was collected from vaccinated individuals after the Sputnik V vaccination and after the Sputnik Light revaccination (Table S1). The blood was centrifuged at 1000g for 10 minutes. The serum was aliquoted and stored at −80°C until use. RBD-specific IgG was determined by ELISA kit «SARS-CoV-2-RBD-ELISA-GAMALEYA» (P3H 2020/10393 2020-05-18).

Before performing the neutralization assay, the serum was thawed and inactivated at +56°C for 30 minutes.

### Virus identification and sequencing

The virus was isolated from a nasopharyngeal swab of a patient who arrived from South Africa. Sample preparation of RNA for sequencing and sequencing were carried out according to the method described earlier [6].

### Cell culture, virus isolation and propagation

Vero E6 cells were used for isolation, initial passage, propagation and titration. Vero E6 cells were maintained in complete Dulbecco’s modified Eagle’s medium (DMEM, HyClone Cytiva, Austria), containing 10% heat-inactivated fetal bovine serum (HI-FBS, Capricorn scientific, Germany), L-glutamine (4 mM, PanEco, Russia), penicillin/streptomycin solution (100 IU/mL; 100 μg/mL, PanEco, Russia) and gentamicin solution (50 μg/mL, Capricorn scientific, Germany).

The SARS-CoV-2 B.1.1.529 virus was isolated from a nasopharyngeal swab. The virus was isolated, propagated and titrated on Vero E6 cells in complete DMEM with 2% HI-FBS. Second passage of virus was harvested, aliquoted and stored at −80°C. The virus was titrated by microtitration method, titers were determined by the 50% tissue culture infective dose (TCID50) method, the titer was determined by Spearman-Kaerber method. All work with infectious SARS-CoV-2 virus was performed under biosafety level 3 (BSL-3) conditions.

### Neutralization assay

Serum NtAb of vaccinated against SARS-CoV-2 variants were analyzed by microneutralization test as described earlier [6]. Briefly, sera samples after inactivation were serially diluted in complete DMEM supplemented with 2% HI-FBS with starting sample dilution at 1:2.5 with two-fold dilution and mixed with 100 TCID50 SARS-CoV-2 at 1:1 ratio (50 μl serum dilution and 50 μl virus suspension) and incubated at 37 °C for 1 h. After that, serum-virus complexes were added to Vero E6 cells in 96-well plates and incubated for 96 h. The cytopathic effect (CPE) was assessed visually, if even a slight damage to the monolayer (1-2 «plaques») was observed in the well, this well was considered as a well with a manifestation of CPE. Neutralization titer was defined as the highest serum dilution without any CPE in two of three replicable wells.

### Statistical analysis

Statistical analysis was performed in GraphPad Prism version 9.2.0 (GraphPad Software Inc., San Diego, CA, USA). For comparison of paired data, Wilcoxon test was used, for comparison of unpaired data – Mann-Whitney test.

## Data Availability

All data produced in the present work are contained in the manuscript.

## Conflict of Interest Disclosures

DIV, DAS, EAS, IFM, GDM, BAG, SDV, TAI, ZOV, LDY and GAL report patents for a Sputnik V immunobiological expression vector, pharmaceutical agent, and its method of use to prevent COVID-19. All other authors declare no competing interests.

## Contributions

LDY - principal investigator. DIV - draft report writing. DIV, GVA, MMM - coordination of the study. PAA, SEV, KNA - PCR and sequencing. SAE, DIV, IAA, KAV (Kovyrshina AV), KAV (Kuzina AV) - virus isolation, propagation, titration. DIV, IAA, KAV (Kovyrshina AV), KAV (Kuzina AV), DAS, EAS, IFM, GDM, BAG, SDV, TAI, ZOV – sera collection, IgG and NtAb determination. DIV, SDV and GVA - data analysis and interpretation. DIV, SDV, GVA, LDY, GAL - the report editing. GAL organized the research and had final responsibility for the decision to submit for publication. All authors critically reviewed the report and approved the final version.

## Supplementary data

**Table S1:** Summary Table of Participants

|  | **Only Sputnik V** | **Sputnik V + Sputnik Light** | **ALL** |
| --- | --- | --- | --- |
| Number of individuals | 9 | 6 | 15 |
| Age, years* | 34<br>(26,5-40,5) | 33<br>(25,8-51,8) | 34<br>(26-42) |
| Months post vaccination<br>or revaccination* | 11<br>(8-11,5) | 2,5 (2,4-2,6) | 8 (2,5-11) |
| Male sex | 3 | 3 | 6 |
| RBD-specific IgG titer** | 1866<br>(1043-3341) | 32254<br>(17809-58415) | 5835<br>(2418-14080) |

\* values are median (interquartile range).
\*\* values are geometric mean (95% confidence interval).

## Notes

### Funding Statement

This study was funded by Russian Direct Investment Fund.

